# Sensorimotor upper limb therapy does not improve somatosensory function and may negatively interfere with motor recovery: a randomized controlled trial in the early rehabilitation phase after stroke

**DOI:** 10.1101/2020.09.15.20194845

**Authors:** De Bruyn Nele, Saenen Leen, Thijs Liselot, Van Gils Annick, Ceulemans Eva, Essers Bea, Lafosse Christophe, Michielsen Marc, Beyens Hilde, Schillebeeckx Fabienne, Alaerts Kaat, Verheyden Geert

**Affiliations:** Department of Rehabilitation Sciences, KU Leuven - University of Leuven, Leuven, Belgium; RevArte - Rehabilitation hospital Antwerp, Antwerp, Belgium; Jessa Hospital Herk-de-Stad, Campus Sint-Ursula, Herk-de-Stad, Belgium; University hospitals Leuven, Department acquired brain injury, Pellenberg, Belgium

**Author notes:** **Correspondence:** Nele De Bruyn.

**Keywords:** Stroke, Upper Extremity, Treatment Outcome, Randomised Controlled Trial, Sensorimotor Therapy

## Abstract

**Question:** Is sensorimotor upper limb (UL) therapy of more benefit for motor and somatosensory outcome than motor therapy?

**Design:** Randomised assessor-blinded multi-centre controlled trial with block randomization stratified for neglect, severity of motor impairment, and type of stroke.

**Participants:** 40 first-ever stroke patients with UL sensorimotor impairments admitted to the rehabilitation centre

**Intervention:** Both groups received 16 hours of additional therapy over four weeks consisting of sensorimotor (N=22) or motor (N=18) UL therapy.

**Outcome measures:** Action Research Arm test (ARAT) as primary outcome, and other motor and somatosensory measures were assessed at baseline, post-intervention and after four weeks follow-up.

**Results:** No significant between-group differences were found for change scores in ARAT or any somatosensory measure between the three time points. For UL impairment (Fugl-Meyer assessment), a significant greater improvement was found for the motor group compared to the sensorimotor group from baseline to post-intervention (mean (SD) improvement 14.65 (2.19) versus 5.99 (2.06); p=0.01) and from baseline to follow-up (17.38 (2.37) versus 6.75 (2.29); p=0.003).

**Conclusion:** UL motor therapy may improve motor impairment more than UL sensorimotor therapy in patients with sensorimotor impairments in the early rehabilitation phase post stroke. For these patients, integrated sensorimotor therapy may not improve somatosensory function and may negatively influence motor recovery.

**Trial registration:** The trial is registered at clinicaltrials.gov NCT03236376.

## 1 Introduction

Somatosensory information is processed when interacting with the environment by touching and manipulating objects. Sensation arising from skin, muscles and joints constitutes the somatosensory ability. Somatosensory function can be divided in three categories. Firstly, the exteroceptive function, consisting of light touch, temperature and pain sensations. Secondly, proprioceptive function existing of position, movement and vibration sense. Lastly, the higher cortical or discriminative function consisting of sharp/dull discrimination, stereognosis and graphesthesia. (1) Somatosensory upper limb (UL) impairment is common after stroke and negatively impacts upon activities of daily living. Approximately 50% of patients encounter somatosensory dysfunction. (2) Differences in prevalence rates are reported for different modalities. Exteroception is impaired in 7-53% of patients, 34-64% encounter proprioceptive deficits and 31-89% have impaired higher cortical function. (3) Moreover, the majority of patients encounter an impairment in more than one modality. A longitudinal study of our research group indicated that in the first week and at six months post stroke, respectively 66% and 28% of the patients with an UL impairment encounter somatosensory impairments in more than one modality, and 50% and 13% in all three modalities. (4)

Somatosensory function is postulated to form an important factor within the motor learning feedforward-feedback mechanism. (5) Lesion studies in animals and humans reported impaired motor control after a focal primary somatosensory cortex (S1) lesion. (6) Motor learning is a key mechanism for stroke recovery. Somatosensory impairments may thus affect motor outcome. A review of Coupar et al. (2012) (7) showed that an intact somatosensory function positively influences motor outcome. More specifically, the presence of somatosensory evoked potentials is reported as a predictor for improved motor recovery. Furthermore, the absence of cortical activation after peripheral somatosensory stimulation is associated with poorer outcome. Clinically, patients with somatosensory impairments are reported to have reduced recovery of dexterity, manipulation skills, grip force regulation and pincer grip. (3) Additionally, longer hospital stay, more social isolation and lower perceived physical activity are reported in patients with somatosensory impairments. (3) Recently, the study of Ingemansson (8) described that proprioceptive impairments at baseline were a negative predictor for treatment outcome even after correction for baseline motor impairment. They reported that 56% of the variation in treatment outcome of robot-assisted finger therapy could be explained by somatosensory system injury together with ipsilesional primary motor cortex (M1) and secondary somatosensory cortex (S2) connectivity.

Somatosensory therapy can improve somatosensory function. Serrada et al. (2019) (9) reported a moderate positive effect for passive somatosensory therapy such as peripheral stimulation, thermal stimulation and intermittent compression therapy. No evidence was presented for active somatosensory therapy due to heterogeneity in outcome measures. However, a positive effect was suggested since all studies reported a positive effect on outcome. The effect of somatosensory therapy on motor function is debated. Grant and colleagues(10) reviewed the effect of somatosensory stimulation on motor performance. They found moderate evidence that somatosensory stimulation does not improve motor performance. Yilmazer and colleagues (2019) (11) on the other hand, showed limited evidence for passive somatosensory therapy and some evidence for the effect of active somatosensory therapy on motor function. Nevertheless, when aiming at improving motor function, a pure somatosensory approach may not be sufficient as it is known that task-specific motor training is effective in improving motor outcome. Due to the coupling between somatosensation and movement in the motor learning mechanism, it may be more beneficial for motor outcome to integrate somatosensory and motor therapy into a sensorimotor approach, than providing motor therapy alone.

The effect of integrated sensorimotor therapy is underinvestigated. De Diego et al. (12) showed a positive effect of sensory stimulation combined with functional activity training in chronic stroke patients on motor and somatosensory function compared to conventional therapy. Similarly, Machackova reported more motor improvement and additional sensory recovery in the group receiving somatosensory stimulation combined with standard motor therapy compared to functional training. (13) Furthermore, afferent stimulation combined with mirror therapy was found to induce greater motor improvements and less synergetic shoulder abduction compared to mirror therapy or functional training only. (14) However, no between-group differences in improvement were reported.

In summary, the important role of somatosensory function for motor performance is well established. Post stroke, the additional effect of somatosensory function and the integration of this function in a sensorimotor therapy program on motor recovery is still poorly understood. Therefore, in this study we compared the effect of a newly developed UL sensorimotor therapy versus motor therapy on UL motor and somatosensory function and functional outcome post stroke. Our research question is ‘Is sensorimotor UL therapy of more benefit for motor and somatosensory outcome than motor therapy?’ We hypothesized integrated sensorimotor therapy to be more beneficial for improving UL function than pure motor therapy.

## 2 Materials and Methods

### 2.1 Design

The methods of our assessor-blinded multicentre randomized controlled trial are described in detail elsewhere. (15) We provide a summary below. This trial is registered at clinicaltrails.gov (NCT03236376) and was approved by the ethical committee of UZ/KU Leuven (s60278). Patients within eight weeks post stroke were randomized (computer-generated) to a four-week additional intervention, based on a block randomisation with type of stroke, presence of neglect and UL motor impairment severity (based on the ability to perform active wrist and finger extension) as stratification factors. We used concealed allocation with opaque envelopes based on an a priori computer generated allocation list with an allocation ratio of 1:1 and stratified blocks of 1, 2 or 3. Allocation was conducted by the principal investigator of the trial, who had no contact with the eligible patients and who was not involved in assessment or therapy provision. The experimental group received 16 hours of additional sensorimotor therapy and the control group received 16 hours of additional motor therapy. Patients were assessed by a blinded assessor at three time points: T1: baseline (pre-intervention) assessment; T2: post-intervention assessment after four weeks of additional therapy; and T3: after four weeks follow-up.

### 2.2 Participants, therapists and centers

First-ever stroke patients were recruited on admission to the rehabilitation ward from four rehabilitation centres in Belgium: UZ Leuven (Pellenberg); Jessa Hospitals (Herk-de-Stad), RevArte (Antwerp) and Heilig Hart Ziekenhuis (Leuven). Inclusion criteria were: first-ever supratentorial stroke within eight weeks post stroke, presence of sensorimotor impairment of the UL based on action research arm test (ARAT) score < 52 out of 57 and a negative composited standardized somatosensory deficit index (16) aged 18 years or older and sufficient cooperation. Patients with musculoskeletal or other neurological disorders, severe communication or cognitive deficits or no informed consent were excluded from this trial.

### 2.3 Intervention

The experimental sensorimotor therapy consisted of 30 minutes of sensory re-learning training based on the SENSe approach(16) and 30 minutes of newly developed sensorimotor training with sensory integrated task-specific motor exercises for the UL, such as sliding over different textures or reaching towards and sorting bottles with a different weight, as described elsewhere. (15) The control ‘motor group’ received 30 minutes of cognitive table-top games with the non-affected UL and 30 minutes of task-specific motor exercises comparable to the sensorimotor exercises, such as sliding over the table or reaching towards the same bottle, but without any emphasis on sensory components. Both groups received 16 one-hour therapy sessions within 4 weeks as an addition to their conventional inpatient therapy program.

### 2.4 Outcome measures

The ARAT, investigating UL activity, was defined as our primary outcome measure. (17) Motor outcome measures included the Fugl-Meyer assessment for the upper extremity (FMA-UE), (18) evaluating motor impairment, and the stroke UL capacity scale (SULCS), (19) assessing functional upper limb use. Somatosensory outcome measures included Erasmus modified Nottingham sensory assessment (Em-NSA) (20) for evaluating exteroception, proprioception and higher cortical functions, perceptual threshold of touch (PTT) (21), assessing light touch perception, texture discrimination test (TDT) (22) for texture discrimination, wrist position sense test (WPST) (23) for proprioceptive discrimination and functional tactile object recognition test (fTORT) (24) to evaluate stereognosis.

### 2.5 Data analysis

Patient characteristics were analysed with descriptive statistics. Normality was checked with the Shapiro-Wilk test (p<0.05). Since all outcome measures were not normally distributed, variables were analysed with counts (percentages) for frequency, and median with interquartile range for ordinal and continuous measures. Between-group differences at baseline were investigated using chi-square or Mann-Whitney U tests. Change scores were calculated between all time points (T2-T1; T3-T2; T3-T1) for experimental and control groups. Effect of treatment group was then investigated with mixed models controlling for age to compare change scores between both groups. Two-tailed p-values, estimated mean differences, and standard error were calculated. Correction for multiple comparison (Bonferroni) was applied and corrected p-value was set at p<0.02.

#### 2.5.1 Secondary analysis

Per protocol, subgroup analysis investigating the effect of therapy group was performed as explained above for patients with mild to moderate, and severe initial motor impairments separately. Subgroups were based on stratification criteria; the ability to perform wrist and finger extension for patients with mild to moderate motor impairments. The a priori power analysis is presented in our protocol. (15)

## 3 Results

### 3.1 Flow of participants, therapists and centers trough the study

A total of 40 stroke patients were recruited with a mean time post stroke of 41 days (SD=13) between September 2017 and October 2019. Of these patients, 22 were allocated to the sensorimotor group and 18 to the motor group. In each group, one patient dropped out from therapy, the first because of medical reasons unrelated to the trial and the second decided to leave the rehabilitation centre. The post-intervention assessment was not performed for two patients because of acute illness in one patient and due to logistic issues in the other patient. The latter did perform the follow-up assessment. Two other individuals, one in each group, were lost to follow-up due to readmission to the acute hospital and because of decline of further participation. No adverse events associated with the interventions were reported. The vast majority of the other patients that were screened were not eligible due to not meeting the criteria ‘first stroke’ or ‘no other neurological or musculoskeletal disorders present affecting the upper limb’.

The flowchart of the study is presented in Figure 1.

**Figure. 1.**
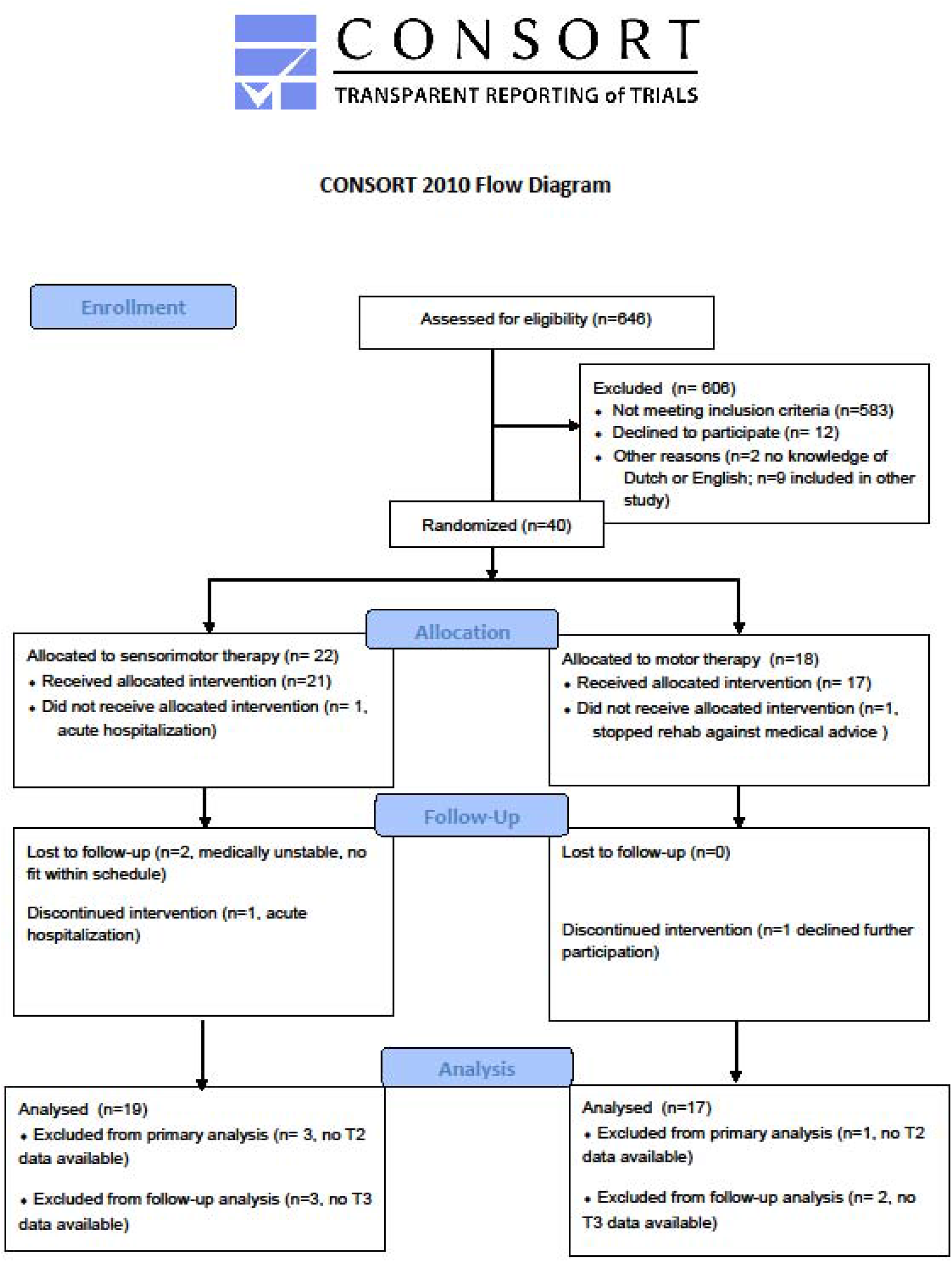
**Flowchart** based on CONSORT guidelines for RCT

Patient characteristics are listed in Table 1 and a lesion overlay map is available in Figure 2. The latter showing a recognizable distribution with common involvement of the middle cerebral artery region. No significant differences were found between groups at baseline, except for age and lateralization; participants in the experimental group were significantly older and had more right hemispheric lesions. Other baseline characteristics were similar for both groups such as time post stroke with a median of 38.5 days (IQR=31-48) for the sensorimotor group and 40 days (IQR=29-54) for the motor group. Baseline performance on the ARAT was 8 points out of 57 (IQR=0-41) for the sensorimotor group and 12 points (IQR=0-35) for the motor group.

**Table 1.**
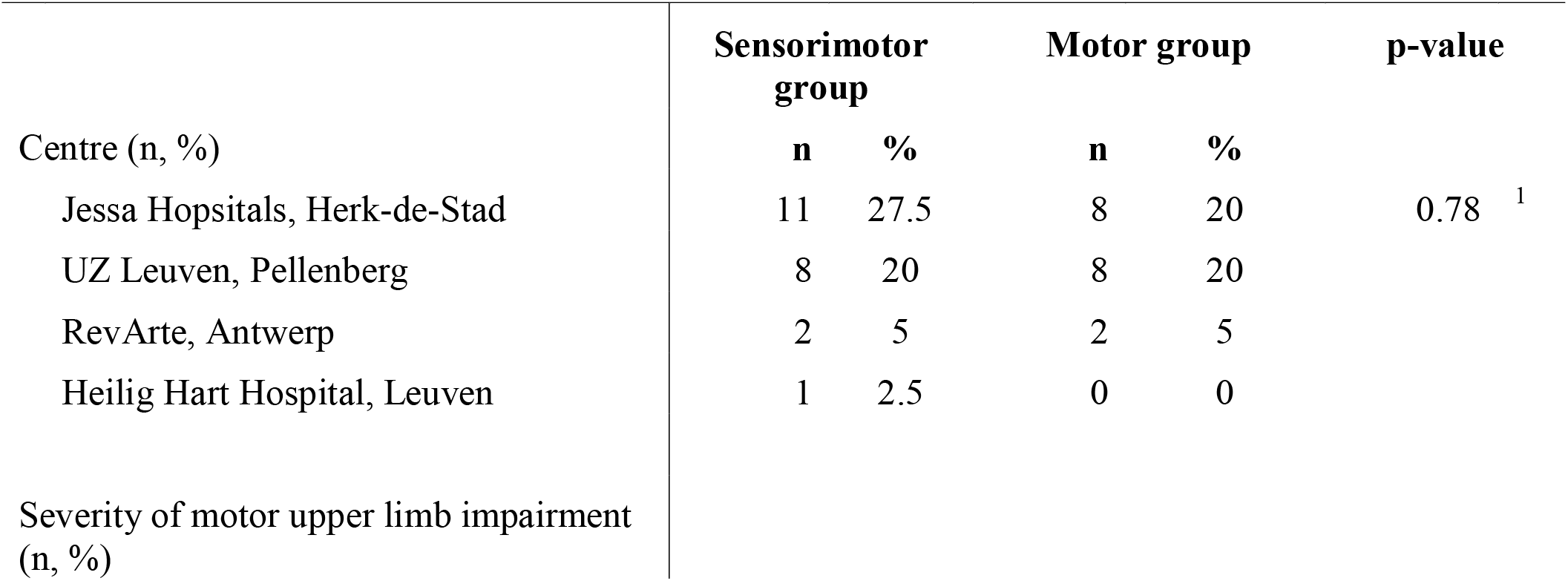
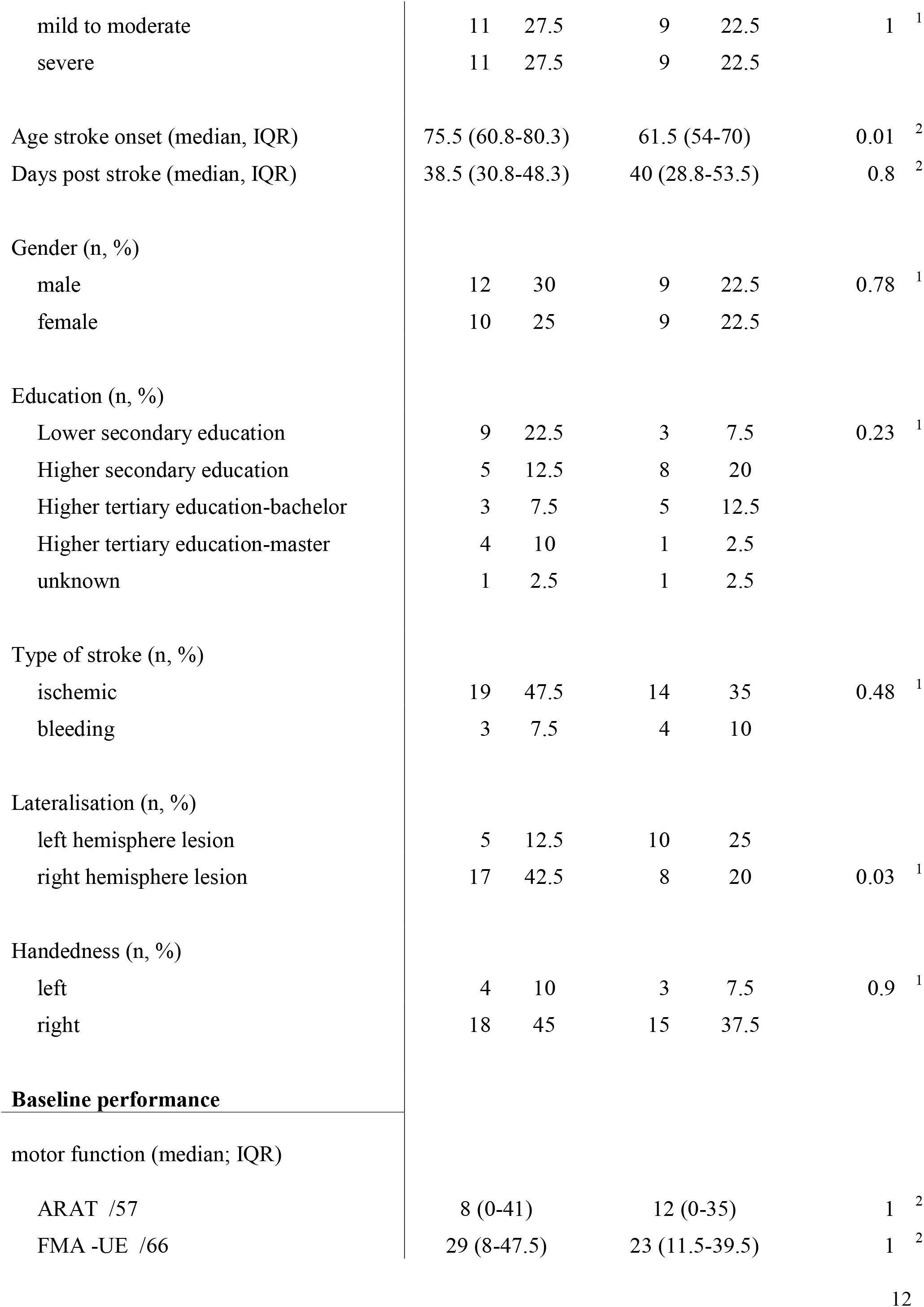
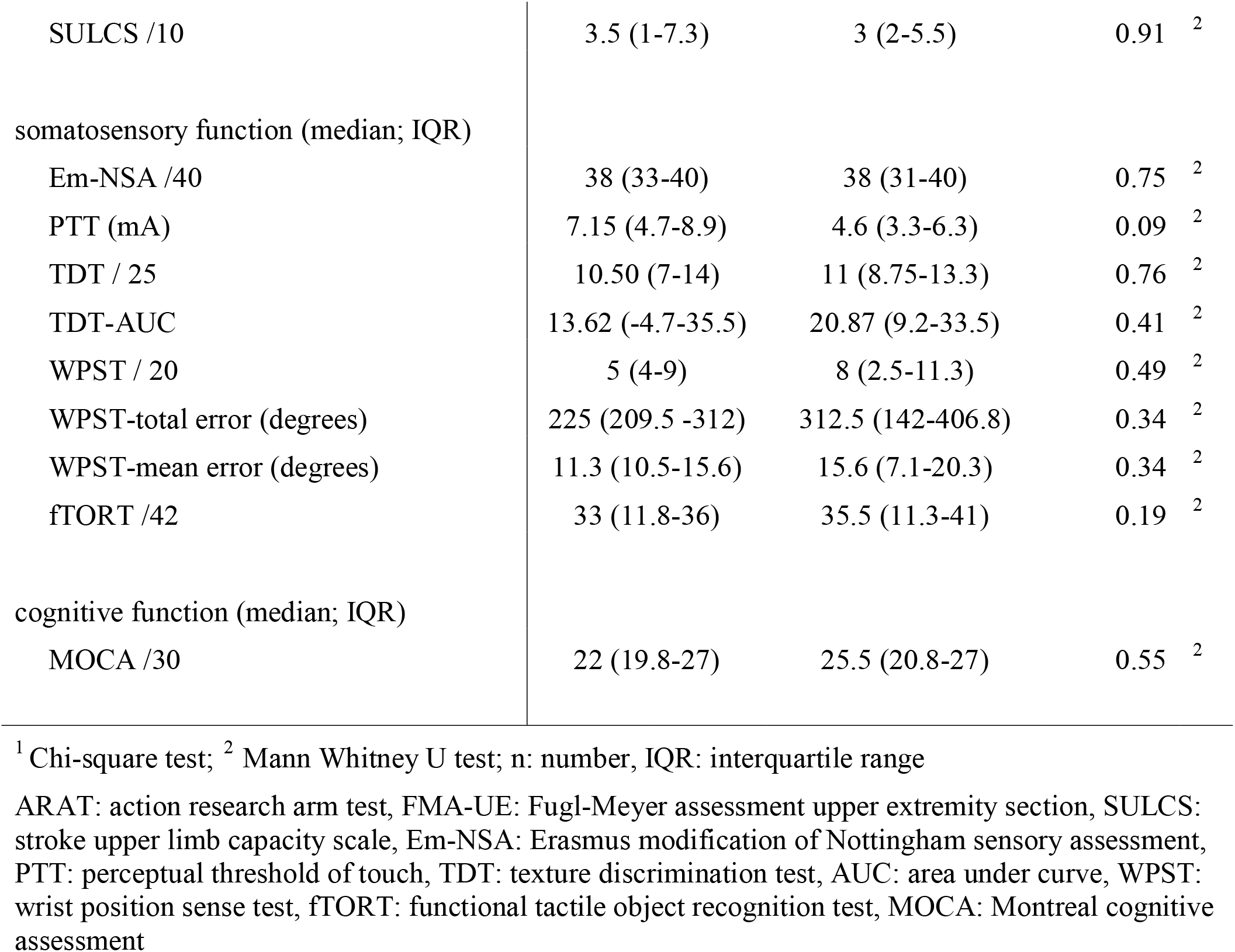
Patient characteristics

|  | Sensorimotor group |  | Motor group |  | p-value |
| --- | --- | --- | --- | --- | --- |
|  | n | % | n | % |  |
| Centre (n, %) |  |  |  |  |  |
| Jessa Hospitals, Herk-de-Stad | 11 | 27.5 | 8 | 20 | 0.78<sup>1</sup> |
| UZ Leuven, Pellenberg | 8 | 20 | 8 | 20 |  |
| RevArte, Antwerp | 2 | 5 | 2 | 5 |  |
| Heilig Hart Hospital, Leuven | 1 | 2.5 | 0 | 0 |  |
| Severity of motor upper limb impairment (n, %) |  |  |  |  |  |

|  |  |  |  |  |  |  |
| --- | --- | --- | --- | --- | --- | --- |
| mild to moderate | 11 | 27.5 | 9 | 22.5 | 1 | <sup>1</sup> |
| severe | 11 | 27.5 | 9 | 22.5 |  |  |
| Age stroke onset (median, IQR) | 75.5 (60.8-80.3) |  | 61.5 (54-70) |  | 0.01 | <sup>2</sup> |
| Days post stroke (median, IQR) | 38.5 (30.8-48.3) |  | 40 (28.8-53.5) |  | 0.8 | <sup>2</sup> |
| Gender (n, %) |  |  |  |  |  |  |
| male | 12 | 30 | 9 | 22.5 | 0.78 | <sup>1</sup> |
| female | 10 | 25 | 9 | 22.5 |  |  |
| Education (n, %) |  |  |  |  |  |  |
| Lower secondary education | 9 | 22.5 | 3 | 7.5 | 0.23 | <sup>1</sup> |
| Higher secondary education | 5 | 12.5 | 8 | 20 |  |  |
| Higher tertiary education-bachelor | 3 | 7.5 | 5 | 12.5 |  |  |
| Higher tertiary education-master | 4 | 10 | 1 | 2.5 |  |  |
| unknown | 1 | 2.5 | 1 | 2.5 |  |  |
| Type of stroke (n, %) |  |  |  |  |  |  |
| ischemic | 19 | 47.5 | 14 | 35 | 0.48 | <sup>1</sup> |
| bleeding | 3 | 7.5 | 4 | 10 |  |  |
| Lateralisation (n, %) |  |  |  |  |  |  |
| left hemisphere lesion | 5 | 12.5 | 10 | 25 |  |  |
| right hemisphere lesion | 17 | 42.5 | 8 | 20 | 0.03 | <sup>1</sup> |
| Handedness (n, %) |  |  |  |  |  |  |
| left | 4 | 10 | 3 | 7.5 | 0.9 | <sup>1</sup> |
| right | 18 | 45 | 15 | 37.5 |  |  |
| **Baseline performance** |  |  |  |  |  |  |
| motor function (median; IQR) |  |  |  |  |  |  |
| ARAT /57 | 8 (0-41) |  | 12 (0-35) |  | 1 | <sup>2</sup> |
| FMA -UE /66 | 29 (8-47.5) |  | 23 (11.5-39.5) |  | 1 | <sup>2</sup> |

|  |  |  |  |
| --- | --- | --- | --- |
| SULCS /10 | 3.5 (1-7.3) | 3 (2-5.5) | 0.91 <sup>2</sup> |
| somatosensory function (median; IQR) |  |  |  |
| Em-NSA /40 | 38 (33-40) | 38 (31-40) | 0.75 <sup>2</sup> |
| PTT (mA) | 7.15 (4.7-8.9) | 4.6 (3.3-6.3) | 0.09 <sup>2</sup> |
| TDT / 25 | 10.50 (7-14) | 11 (8.75-13.3) | 0.76 <sup>2</sup> |
| TDT-AUC | 13.62 (-4.7-35.5) | 20.87 (9.2-33.5) | 0.41 <sup>2</sup> |
| WPST / 20 | 5 (4-9) | 8 (2.5-11.3) | 0.49 <sup>2</sup> |
| WPST-total error (degrees) | 225 (209.5 -312) | 312.5 (142-406.8) | 0.34 <sup>2</sup> |
| WPST-mean error (degrees) | 11.3 (10.5-15.6) | 15.6 (7.1-20.3) | 0.34 <sup>2</sup> |
| fTORT /42 | 33 (11.8-36) | 35.5 (11.3-41) | 0.19 <sup>2</sup> |
| cognitive function (median; IQR) |  |  |  |
| MOCA /30 | 22 (19.8-27) | 25.5 (20.8-27) | 0.55 <sup>2</sup> |

<sup>1</sup> Chi-square test; <sup>2</sup> Mann Whitney U test; n: number, IQR: interquartile range
ARAT: action research arm test, FMA-UE: Fugl-Meyer assessment upper extremity section, SULCS: stroke upper limb capacity scale, Em-NSA: Erasmus modification of Nottingham sensory assessment, PTT: perceptual threshold of touch, TDT: texture discrimination test, AUC: area under curve, WPST: wrist position sense test, fTORT: functional tactile object recognition test, MOCA: Montreal cognitive assessment

**Figure 2.**
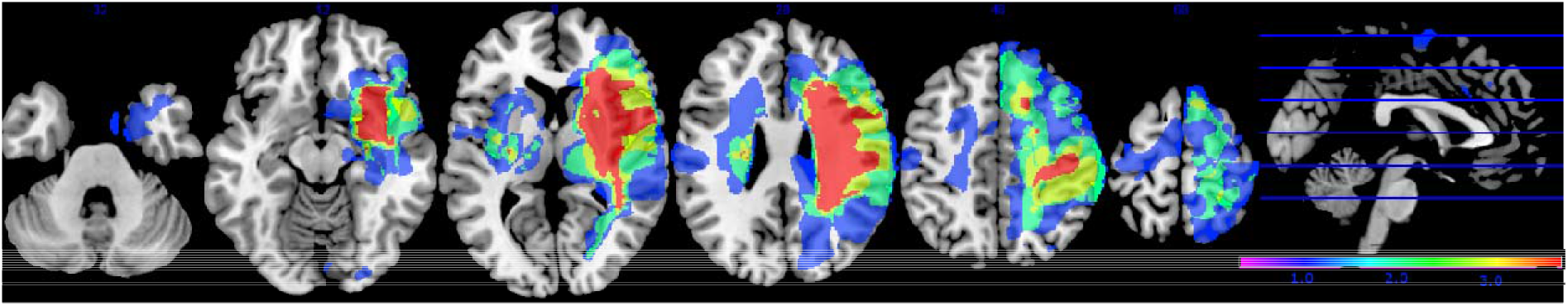
**Lesion overlay map** of stroke lesion location of patients with available magnetic resonance imaging (MRI) scan (n=30). Colour indicates increasing number of patients with inclusion of that voxel into the lesion from blue to red (low number: blue; high number: red).

### 3.2 Between-group intervention effect

Results of between-group comparisons are presented in Table 2 and Figure 3. A trend towards between-group difference in favour of the motor group was found for our primary outcome measure (ARAT) in changes between all time points. From baseline to post-intervention, a significant greater improvement was found for the motor group in comparison to the sensorimotor group for FMA-UE, and a similar trend was found for SULCS. For motor impairment (FMA-UE), mean pre-to-post improvement in the motor group was 14.65 points, compared to 5.99 points in the sensorimotor group, resulting in a mean difference in improvement (age-adjusted) in favour of the motor group of 8.66 points (standard error (SE) 3.12, t=-2.77, p=0.01). From pre-intervention to follow-up, the significant greater improvement for the motor group in comparison to the sensorimotor group for FMA-UE and a trend for SULCS were retained. For motor impairment (FMA-UE), age-adjusted mean difference in favour of the motor group was 10.63 points (SE=3.39, t= −3.14, p=0.003). No significant between-group difference in changes between time points was found for any of our secondary somatosensory measures. Individual delta changes over time and individual time courses of motor recovery can be found in supplementary figure 1 and 2 respectively.

**Table 2.** Between group comparison of an intervention effect corrected for age

| Motor function |  |  |  |  |  |  |  |
| --- | --- | --- | --- | --- | --- | --- | --- |
|  |  | ARAT /57 | FMA /66 | SULCS /10 | ABILHAND (logits) |  |  |
| T2-T1 | Sensorimotor group | 4.87 (2.32) | 5.99 (2.06) | 0.84 (0.38) | 4.86(1.42) |  |  |
|  | Motor group | 11.06 (2.46) | 14.65 (2.19) | 1.87 (0.37) | 4.13 (1.47) |  |  |
|  | p-value | 0.08 | 0.01* | 0.08 | 0.74 |  |  |
|  | 95%CI | (-13.24-0.87) | (-15.03--2.29) | (-2.16-0.11) | (-3.79 -5.24) |  |  |
| T3-T2 | Sensorimotor group | 2.00(1.42) | 1.20 (1.65) | 0.28 (0.37) | 5.00(1.55) |  |  |
|  | Motor group | 6.07 (1.53) | 2.98 (1.72) | 0.75 (0.39) | 0.28 (1.71) |  |  |
|  | p-value | 0.08 | 0.48 | 0.41 | 0.06 |  |  |
|  | 95%CI | (-8.67-0.46) | (-6.92-3.35) | (-1.61-0.68) | (-0.28-9.68) |  |  |
| T3-T1 | Sensorimotor group | 7.65 (2.73) | 6.75 (2.29) | 0.90 (0.45) | 9.12 (1.67) |  |  |
|  | Motor group | 16.32 (2.98) | 17.38 (2.37) | 2.60 (0.45) | 4.26 (1.76) |  |  |
|  | p-value | 0.04 | 0.003* | 0.02 | 0.07 |  |  |
|  | 95%CI | (-17.00--0.34) | (-17.50--3.76) | (-3.04--0.36) | (-0.32-10.05) |  |  |
| Somatosensory function |  |  |  |  |  |  |  |
|  |  | Em-NSA /40 | PTT /10mA | TDT-AUC | WPST total error degrees | WPST mean error degrees | ftORT /42 |
| T2-T1 | Sensorimotor group | 1.48(1.37) | -1.15 (0.54) | 10.18 (6.13) | -56.43 (28.50) | -1.83 (1.32) | 2.41 (1.36) |
|  | Motor group | 2.01 (1.36) | -0.15 (0.57) | 5.11 (6.41) | -62.08 (30.88) | -3.51 (1.41) | 3.39 (1.43) |
|  | p-value | 0.79 | 0.22 | 0.58 | 0.90 | 0.40 | 0.63 |
|  | 95%CI | (-4.65-3.57) | (-2.62 -0.62) | (-13.53-23.65) | (-83-94) | (-2.35-5.70) | (-6.02-2.65) |
| **T3-T2** | Sensorimotor group | 0.85 (1.09) | -0.41 (0.39) | 0.22 (4.53) | -7.61 (22.22) | -0.07 (1.08) | 0.08 (0.99) |
|  | Motor group | 1.31 (1.22) | -0.37 (0.44) | -2.14 (4.89) | -13.54 (24.00) | -0.90 (1.15) | 0.40 (1.06) |
|  | p-value | 0.79 | 0.94 | 0.79 | 0.87 | 0.62 | 0.83 |
|  | 95%CI | (-3.94-3.03) | (-1.30 -1.20) | (-12.46-16.29) | (-65-77) | (-2.50 -4.16) | (-3.41 -2.75) |
| **T3-T1** | Sensorimotor group | 1.57 (1.47) | -1.39 (0.44) | 10.05 (7.00) | -33.13 (32.30) | -1.83 (1.32) | 2.41 (1.43) |
|  | Motor group | 3.66 (1.49) | -1.14 (0.50) | 2.45 (7.52) | -94.46 (34.65) | -3.51 (1.41) | 4.09 (1.53) |
|  | p-value | 0.34 | 0.73 | 0.48 | 0.22 | 0.40 | 0.44 |
|  | 95%CI | (-6.49-2.32) | (-1.63-1.15) | (-13.75-8.94) | (-37 -160) | (-2.35-5.70) | (-6.02-2.65) |

Estimated mean and standard error of changes scores (T2-T1, T3-T2, T3-T1) are presented for both groups; p-values based on mixed models with age stroke onset (years) as covariate to evaluate differences between the change scores of both groups. Correction for multiple comparison was set on $p < 0.02$ . ARAT: action research arm test, FMA-UE: Fugl- Meyer assessment upper extremity section, SULCS: stroke upper limb capacity scale, Em-NSA: Erasmus modification of Nottingham sensory assessment, PTT: perceptual threshold of touch, TDT: texture discrimination test, AUC: area under curve, WPST: wrist position sense test, fTORT: functional tactile object recognition test

**Figure 3.**
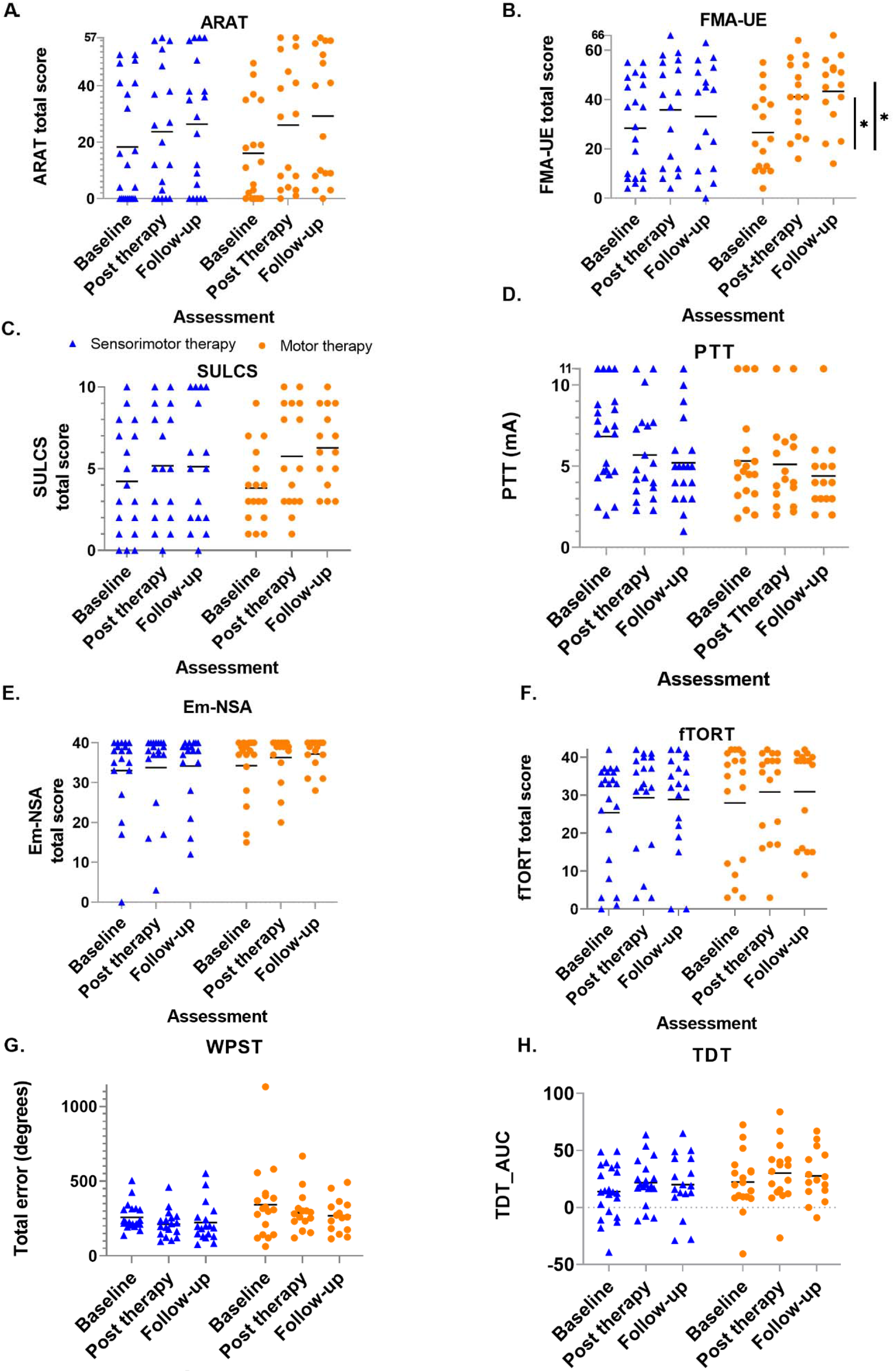
**Scatterplot of outcome variables** for each group at each time point – every dot (motor therapy) or triangle (sensorimotor therapy) at one time point represents the raw value of a patient; raw median scores indicated with horizontal bar. Vertical bars indicate significant differences in change scores between both groups for * *p=0.01; ARAT: Action Research Arm Test, FMA-UE: Fugl-Meyer assessment upper extremity part, SULCS stroke upper limb capacity scale, Em-NSA: Erasmus modification of Nottingham Sensory Assessment; PTT: perceptual threshold of touch (mA), TDT_AUC: texture discrimination test area under curve score, WPST: wrist position sense test mean error (degrees), fTORT functional tactile object recognition test

#### 3.2.1 Secondary analysis

Similar results were found for the subgroup of patients with initial severe motor UL impairments (see Table 3 in eAddenda). A trend of between group differences towards higher change scores was found for the motor group from baseline to post-intervention for FMA-UE, and ARAT; from post-intervention to follow-up for SULCS and ARAT; and from baseline to follow-up for FMA-UE and ARAT. Significant higher change scores were found for the motor group from baseline to follow-up for SULCS. Patients with initial mild to moderate motor UL impairments showed significant higher change scores in the motor group for SULCS from baseline to follow-up. Trends towards significant higher change scores were found for FMA-UE in the motor group and for PTT in the sensorimotor group.

## 4 Discussion

In this study, we compared the effect of a newly developed UL sensorimotor therapy versus motor therapy on UL motor and somatosensory function and functional outcome in the early rehabilitation phase post stroke. In contrast to our hypothesis, we could not show a beneficial effect of sensorimotor therapy. Moreover, the results suggest a better improvement in UL motor impairment from baseline to post-intervention and to follow-up assessment for the motor group.

These results are surprising in that we assumed that the integration of a somatosensory component in a motor therapy approach would improve motor recovery due to sensorimotor coupling and the importance of somatosensory function for motor learning. (2) A number of reasons for this finding that contrasts our hypothesis could be considered. First, the sensorimotor group was older compared to the motor group and it is known that age is a predictor of stroke outcome. (25) Thus, we corrected for age in our analysis but still found significant between-group differences. Second, we conducted a dose-matched trial for additional therapy time, which, for both groups, was two times 30 minutes per day. However, number of repetitions could be different between both groups with a higher number of repetitions for the motor group. This could be explained by the nature of the exercises. In the sensorimotor group, patients were asked to focus on the somatosensory input during motor execution, which could have reduced the number of movements performed. Additionally, this focus on somatosensory input could change the prioritization of attention towards the somatosensory task inducing mutual interference. This mutual interference is characterized by a deterioration of performance of both tasks. (26, 27) Further, within the motor learning literature, dual task training consisting of a motor task with a cognitive task has shown to improve motor performance less effectively than motor task training on his own. (28, 29) However, after a motor-cognitive training, performance on this motor-cognitive task is improved but performance of the single motor task is still at the baseline level. (30) This mechanism of context and task specific improvements could be an explanation of our findings in favour of the motor group, which were tested into the same context and task as practiced during therapy. So it could be that we were not able to measure the improvements of the sensorimotor therapy group since we were not able to measure the improvement in integration of somatosensory and motor function due to the lack of assessment method available. To further elaborate on the cognitive sensorimotor interference hypothesis, differences between therapy groups could exist of additional somatosensory integration task for the sensorimotor group, which could lead to cognitive sensorimotor interference with prioritization of the sensory input. In healthy adults, these kinds of daily life movements such as reaching towards a cup or sliding over a surface are automated and thus allow the person to divide the attention towards other (sensory) input without any influence on motor performance. However, in stroke patients, high attention levels are needed to perform even simple sliding or reaching exercises. (26) The addition of somatosensory input could thus induce an allocation of the attention towards the sensory input resulting in impaired performance of the primary (motor) movements. Hence, the integration of a clinical somatosensory component into motor therapy may not be of added value for motor recovery in the early rehabilitation phase. Further research, implementing and evaluating the effect of a revised sensorimotor therapy approach, is needed to provide better insight in effective sensorimotor therapy models and the long-term effects of sensorimotor therapy.

Another result of our trial is that somatosensory function may not improve differently between groups. Only a trend towards better light touch improvement in the sensorimotor group for patients with initial mild to moderate UL impairments was found. We hypothesized the group receiving sensorimotor therapy to have greater improvements of somatosensory function, compared to the pure motor group, since the former received 30 minutes of specific somatosensory training based on the SENSe approach. (16) In contrast to other somatosensory and sensorimotor therapy stroke trials, we could not find differences in improvements in somatosensory function. Differences in methodology exist between our trial and previous studies and could have influenced the results, such as phase post stroke (subacute in our trial versus chronic in earlier work), initial UL somatosensory or motor impairment severity and time spent per session (30 minutes in our trial compared to 60-90 minutes). However, a similar amount of total training time was provided. The majority of other trials in this domain have focused on patients with chronic stroke (12, 14, 16, 31) recruiting people with persistent somatosensory impairments. Initial rather mild somatosensory impairments of our sample could further explain the lack of between-group differences. Only PTT showed a trend towards between-group differences within the initial mild to moderately impaired patients. Biological recovery could explain the overall lack of difference between groups. Proportional recovery of exteroceptive and proprioceptive somatosensory function is reported to be higher than motor recovery.(32) Additionally, most studies focused on patients with initial mild to moderate motor impairments, (13, 14, 31) which allows the patient to divide the attention more towards other (sensory) input, with less influence on motor performance. Last, all studies except one provided somatosensory stimulation in addition to motor training, without implementing the integration of both. Only Borstad et al. included integration of somatosensory and motor function by performing sorting exercises with different features such as weight or size. (31) This study only reported two cases and did not include control participants. Interestingly and similar to our study, they did find improvements in motor, but not somatosensory function for one of both cases. Furthermore, our findings are in line with the systematic review by Grant and colleagues who concluded that there is low to moderate quality evidence that somatosensory stimulation does not improve motor function, impairment or UL activity. (10)

This trial was preregistered at clinicaltrails.gov (NCT03236376) and the protocol was published. (15) Some adaptations occurred after registration and publication. First, predetermined sample size was not reached due to slow recruitment. Therefore, we included two additional rehabilitation centres but we were only able to include a limited number of participants from these centres. Thus, our study should be considered a phase II trial. Second, the nine hole peg test was not included in further analysis since too many patients were not able to perform this test due to severity of upper limb impairment. Third, subgroup analysis based on level of cognition is not reported, since no difference was found during between group analysis. Subgroup analysis based on the presence of neglect was also not performed since only three patients had visuospatial neglect based on the star cancellation test. Results of brain imaging analysis as described in the protocol paper will be published in a separate report.

This study has some limitations. First, the sample size is rather small and leads to reduced power of the study, but comparable with phase II studies in the field. (33-36) Results should be interpreted with caution due to the large confidence intervals but the higher between-group motor improvement for motor therapy could be considered substantial and clinically relevant. Hence, our trial may inform necessary further studies in this domain. Second, the follow-up was only four weeks after additional therapy, which may be too short to find retention effects. Evaluation after three or six months could reveal interesting insights in the long-term effect of therapy. Third, blinding of the assessor was not always possible due to reactions of the patients. Certainly, patients who received sensorimotor therapy could react on the assessment with a response of recognition. However, the assessor was instructed to not pay attention to these reactions. Fifth, concealment was done by the principle investigator, who was not involved in clinical assessment or therapy provision. However, it could be preferred to involve an independent person for this aspect of the methodology. Last, hours of conventional therapy were registered, however the content of this therapy was not. On the other hand, a good balance was obtained in number of patients with mild to moderate or severe impairments receiving both therapy approaches over the different centers. The additional therapy was provided by one therapist in all centers to offer standardization.

To conclude, our results suggest that motor therapy may improve UL motor function to a greater extent compared to sensorimotor therapy in the early rehabilitation phase post stroke for patients with sensorimotor upper limb deficits. Further research is warranted, to investigate whether patients with sensorimotor UL deficits benefit from integrated sensorimotor training and how this training can be delivered effectively.

## Data Availability

data is available from the corresponding author upon reasonable request

## 5 Acknowledgments

We would like to acknowledge the contribution of all physiotherapists, occupational therapists and nursing staff of the neurorehabilitation wards of UZ Leuven, campus Pellenberg; Jessa hospitals Herk-de-Stad; Heilig Hart Hospital Leuven; and RevArte Antwerp. More specifically, we would like to acknowledge Anne-Leen Hindrikx, Elke Janssens, Peter Popelier, Margreet Van Dijk, Bea Slaets, Walter Habils, Sabine Vermeiren and Karen Deceuninck who assisted with recruitment and scheduling of therapy and assessment. Clara De Somer is acknowledged for her support within the research project. Finally, are grateful to all patients who participated in this trial. This work was supported by Flanders Research Fund (FWO) (1189819N and 1519719N).

## 11 Supplementary Material

**Supplementary table 1.**
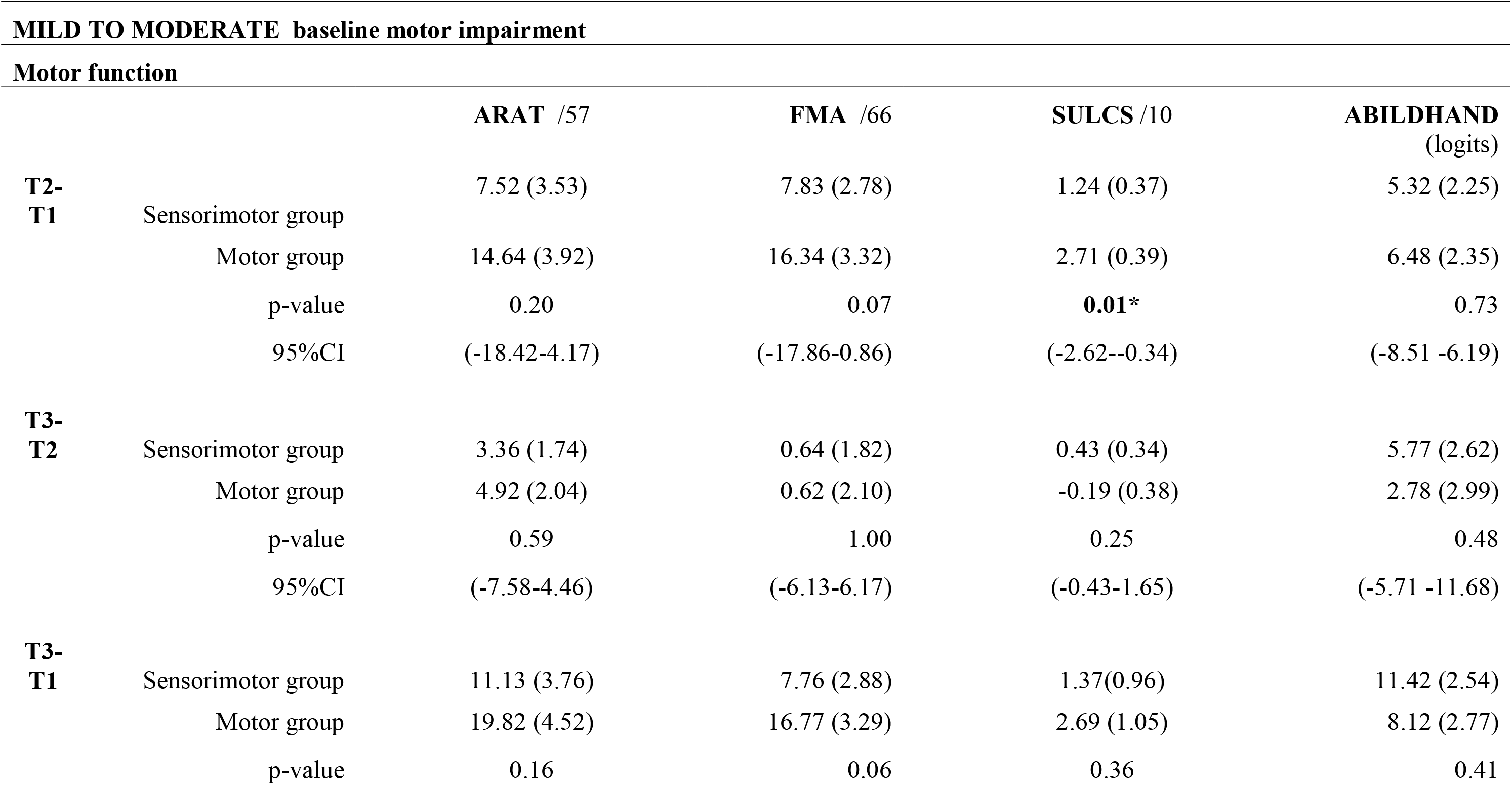
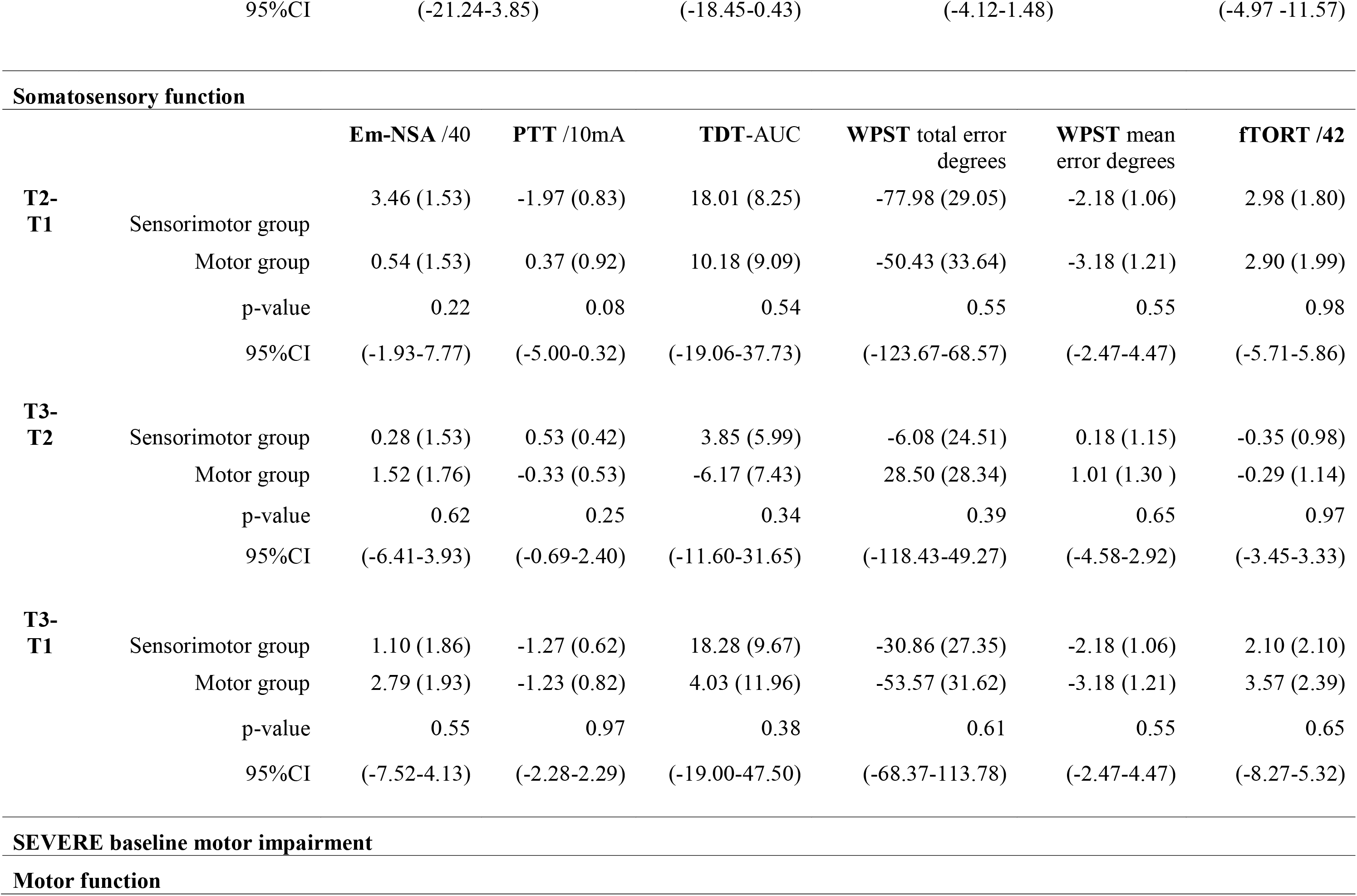
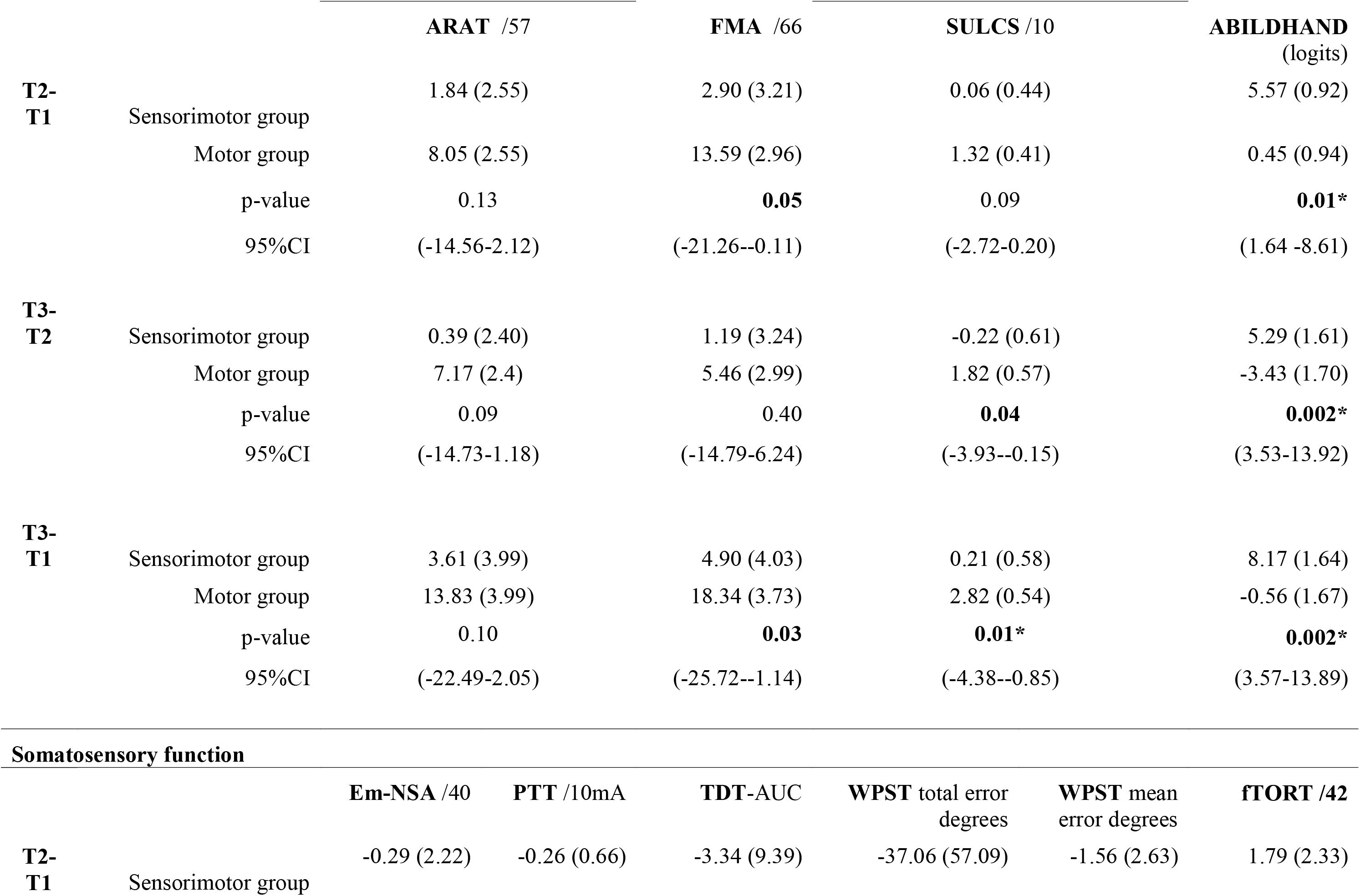
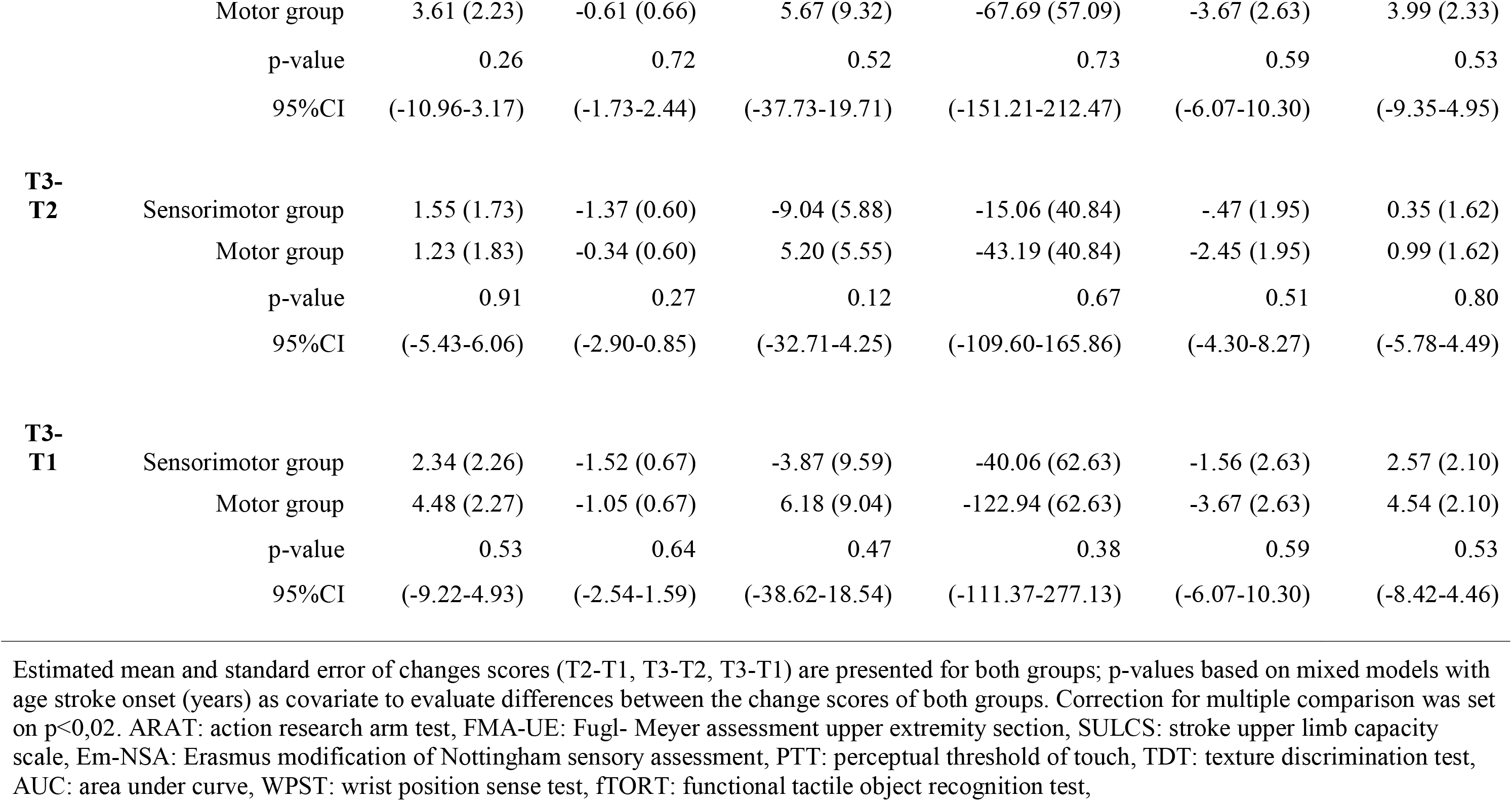
Subgroup analysis of between group comparison of an intervention effect corrected for age.

**Supplementary Figure 1.**
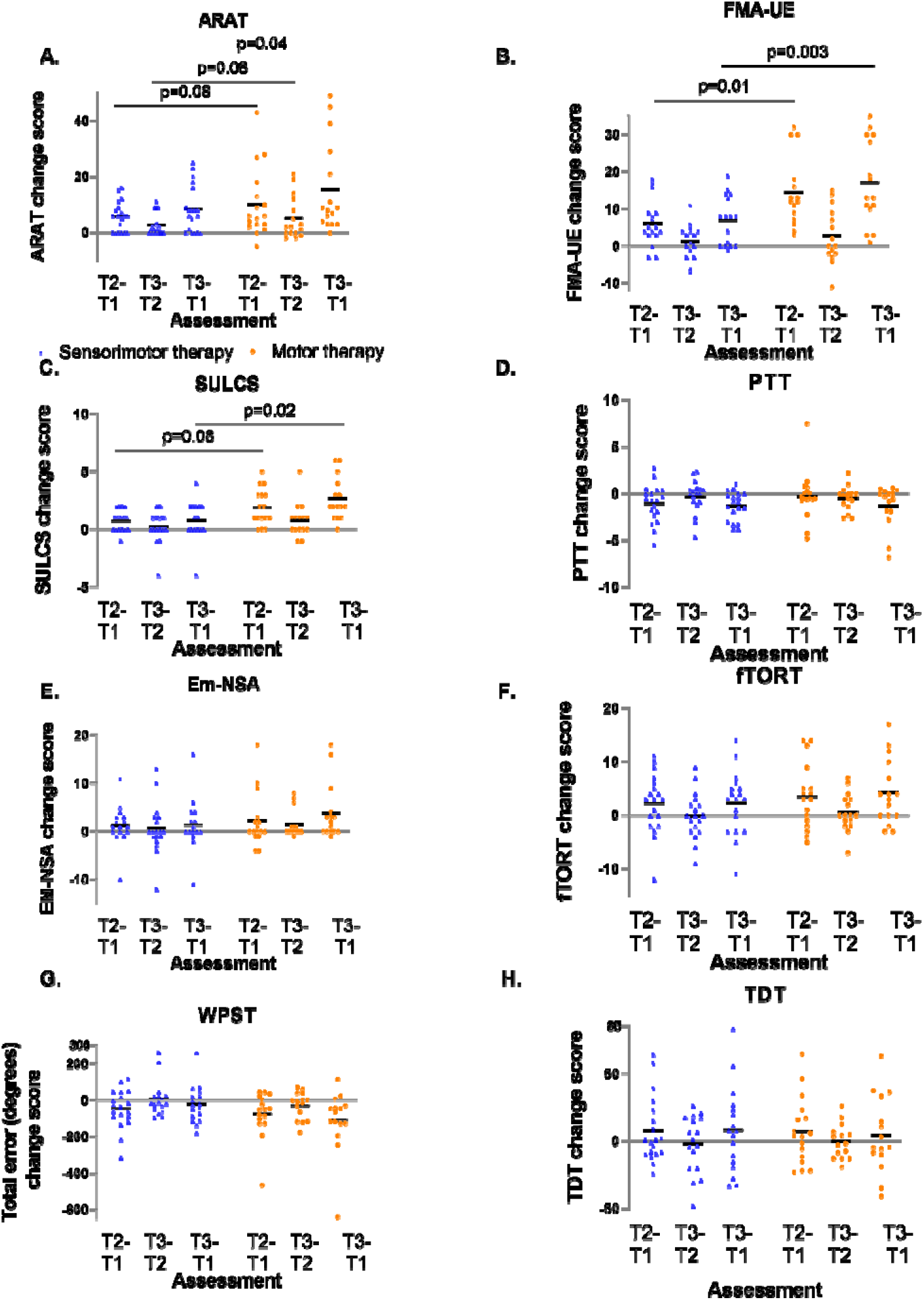
Individual delta changes over time. every dot (motor therapy) or triangle (sensorimotor therapy) at one time point represents the delta change score of a patient; raw median scores indicated with horizontal bar. ARAT: Action Research Arm Test, FMA-UE: Fugl-Meyer assessment upper extremity part, SULCS stroke upper limb capacity scale, Em-NSA: Erasmus modification of Nottingham Sensory Assessment; PTT: perceptual threshold of touch (mA), TDT_AUC: texture discrimination test area under curve score, WPST: wrist position sense test mean error (degrees), fTORT functional tactile object recognition test

**Supplementary Figure 2.**
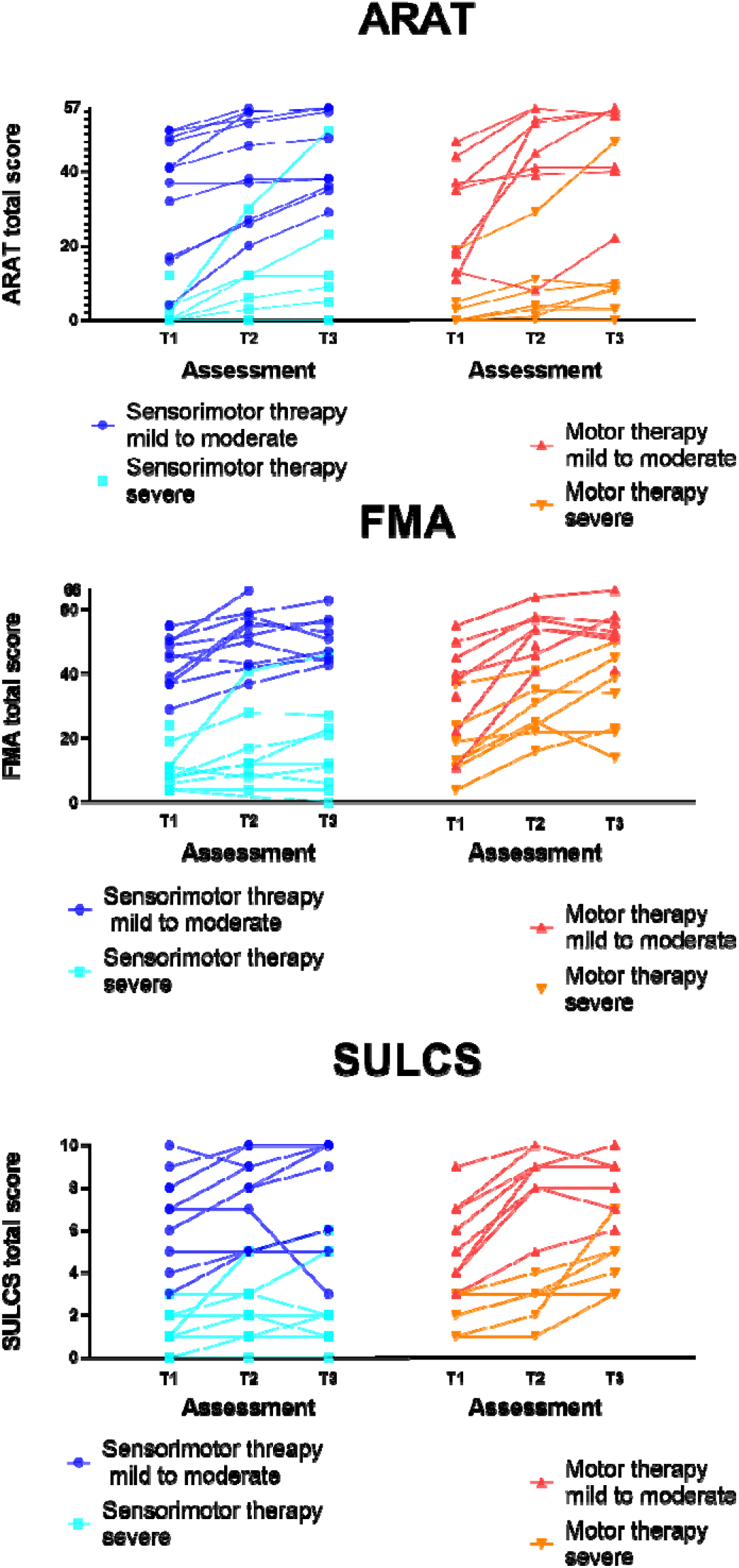
Individual time courses of motor recovery. Every dot/square (sensorimotor therapy) or triangle (motor therapy) at one time point represents the raw value of a patient. Subdivision is made for patients with mild to moderate and severe baseline motor impairments. ARAT: Action Research Arm Test, FMA-UE: Fugl-Meyer assessment upper extremity part, SULCS: stroke upper limb capacity scale

